# Epidemiological and Economic Impact of COVID-19 in the US

**DOI:** 10.1101/2020.11.28.20239517

**Authors:** Jiangzhuo Chen, Anil Vullikanti, Joost Santos, Srinivasan Venkatramanan, Stefan Hoops, Henning Mortveit, Bryan Lewis, Wen You, Stephen Eubank, Madhav Marathe, Chris Barrett, Achla Marathe

**Affiliations:** Network Systems Science and Advanced Computing Division, Biocomplexity Institute, University of Virginia, Charlottesville, VA 22904, USA; Department of Computer Science, University of Virginia; Department of Engineering Management and Systems Engineering, George Washington University, Washington, DC 20052, USA; Department of Engineering Systems and Environment, University of Virginia; Department of Public Health Sciences, University of Virginia

## Abstract

This research measures the epidemiological and economic impact of COVID-19 spread in the US under different mitigation scenarios, comprising of non-pharmaceutical interventions. A detailed disease model of COVID-19 is combined with a model of the US economy to estimate the direct impact of labor supply shock to each sector arising from morbidity, mortality, and lockdown, as well as the indirect impact caused by the interdependencies between sectors. During a lockdown, estimates of jobs that are workable from home in each sector are used to modify the shock to labor supply. Results show trade-offs between economic losses, and lives saved and infections averted are non-linear in compliance to social distancing and the duration of lockdown. Sectors that are worst hit are not the labor-intensive sectors such as Agriculture and Construction, but the ones with high valued jobs such as Professional Services, even after the teleworkability of jobs is accounted for. Additionally, the findings show that a low compliance to interventions can be overcome by a longer shutdown period and vice versa to arrive at similar epidemiological impact but their net effect on economic loss depends on the interplay between the marginal gains from averting infections and deaths, versus the marginal loss from having healthy workers stay at home during the shutdown.

## 1 Introduction

According to the Bureau of Labor Statistics, the US unemployment rate in October 2020 stands at 6.9% and the number of unemployed at 11.1 million. This is likely an underestimated number since it does not include individuals who have stopped looking for employment [16] due to poor economic prospects. Even though both measures have declined for 6 months consecutively, the unemployment rate is still higher by 3.5% and the number of unemployed by 5.3 million, compared to pre-COVID-19 levels in February 2020. The US economy shrank by an annual rate of 4.8% in the first quarter of 2020 and by a shocking 32.9% in the second quarter, which has been the largest drop seen since 1945. The number of COVID-19 cases have crossed 12 million and number of deaths over 258,000 in November 2020 [31].

This research builds a comprehensive system that combines the epidemiological model developed to study the spread of COVID-19 with a detailed model of the US economy to understand a sector wise economic impact from a shock to labor supply caused by the pandemic. Note that the focus of this paper is only on the shock encountered by the economic sectors from the supply side, and not the demand side which has also dropped due to the high unemployment rate and a bleak economic outlook. We consider a number of counterfactual scenarios that comprise of various social distancing measures such as the stay-home order, voluntary home isolation of the symptomatic individuals, and school closure. We measure economic losses from the drop in labor supply in each sector due to stay-home order, absenteeism due to illness and deaths, cascading loss to/from other sectors due to interdependencies between sectors, and the economic burden caused by the medical treatment of the infected. We vary compliance to interventions and duration of the stay-home order to determine their impact on economic and epidemiological outcomes and the trade-offs between them.

This research is an extension of the work done in [15] which only focused on estimating the medical cost of treatment for COVID-19 cases under the same mitigation scenarios and the disease model. Here we calculate overall economic losses from a societal perspective which include the medical cost of illness, cost of intervention or social distancing i.e. healthy individuals unable to go to work, direct loss in productivity due to morbidity and mortality of workers, and the indirect loss caused by the interdependencies between sectors. We also estimate the effect of intervention scenarios on cases and deaths averted in the US.

This level of detailed analysis has not been done in the literature and can provide guidance to public health officials for developing strategies to balance the emergence of infections and deaths with the economic costs of the social distancing strategies. A longer duration of stay-home order causes economic losses even after accounting for telework, but it also significantly reduces infections and deaths, and losses caused by the medical treatment of the infected.

## 2 Related Work

There have been several papers that study the economic impact of COVID-19. Eichenbaum et al. [21] study the interaction between economic decisions and epidemic outcomes and find that the competitive equilibrium is not socially optimal because infected people do not fully internalize the effect of their economic decisions on the spread of the virus. Their results show that an optimal containment strategy that starts early and ramps up with infections, can cause a large recession but save about half a million lives, assuming no treatment or vaccines are available.

Work by [1] shows that differential targeting of risk/age groups outperform uniform social distancing policies. Most of the economic gains in this study are realized from implementing stricter lockdown policies on the oldest age group. However a fully targeted policy can be challenging to implement and ethically questionable.

Baker et al. [4] characterize the uncertainty using stock market volatility measures, newspaper-based measures of uncertainty and survey-based perceptions of business level uncertainty; and find that more than half of the contraction in US economy is caused by COVID-induced uncertainty.

Toda (2020) [38] uses an SIR model to study the impact of the epidemic on the stock market. Jones et al. [25] use an SIR contagion model and a model for consumption and production to analyze optimal mitigation policies and interactions between economic activity and epidemic dynamics.

They discuss congestion externality i.e. when hospital capacity is exceeded, the risk of death becomes higher but agents do not internalize the impact of their decisions on others and therefore behave in a socially sub-optimal way. Other papers that study the economic impact of COVID-19 and pandemics are [7, 10, 17, 37].

The ripple effects of pandemics across a regional economy are studied using an input-output model in [32, 35]. The impacts of pandemic-induced workforce disruptions are assessed using economic loss as well as inoperability, which measures the extent to which sectors are unable to produce their ideal level of output.

The novelty of our research lies in building a detailed integrated system that combines a network based population model with an epidemiological model and an economic model. The disease spreads on the social network, as determined by the COVID-19 disease model; non-pharmaceutical interventions remove particular edges in the social network depending upon the type of interventions and compliance rate; the duration of the interventions determine the length of the time edges are removed for; the outcome of the spread is captured in terms of infections and deaths, which determine the shock to labor-supply in specific economic sectors, as determined by the occupation of individuals who are sick or dead, as well as those who are healthy but unable to work due to a lockdown. These shocks as well as the interdependencies between the sectors determine the sectoral and overall economic impact.

## 3 Data and Methods

This research integrates a variety of datasets to build a comprehensive model that includes individuals, their interactions, their health states over time as the disease spreads over the social contact network, their behaviors in terms of compliance with interventions and its effect on their health states, and the impact of their health outcomes on each economic sectors’ labor supply and hence sectors’ output. Figure 1 shows the overall systems level architecture of the modeling framework, its various components and how they are linked together. Below we describe the various datasets and models used in this framework and how they have been synthesized to build an integrated system.

**Figure 1:**
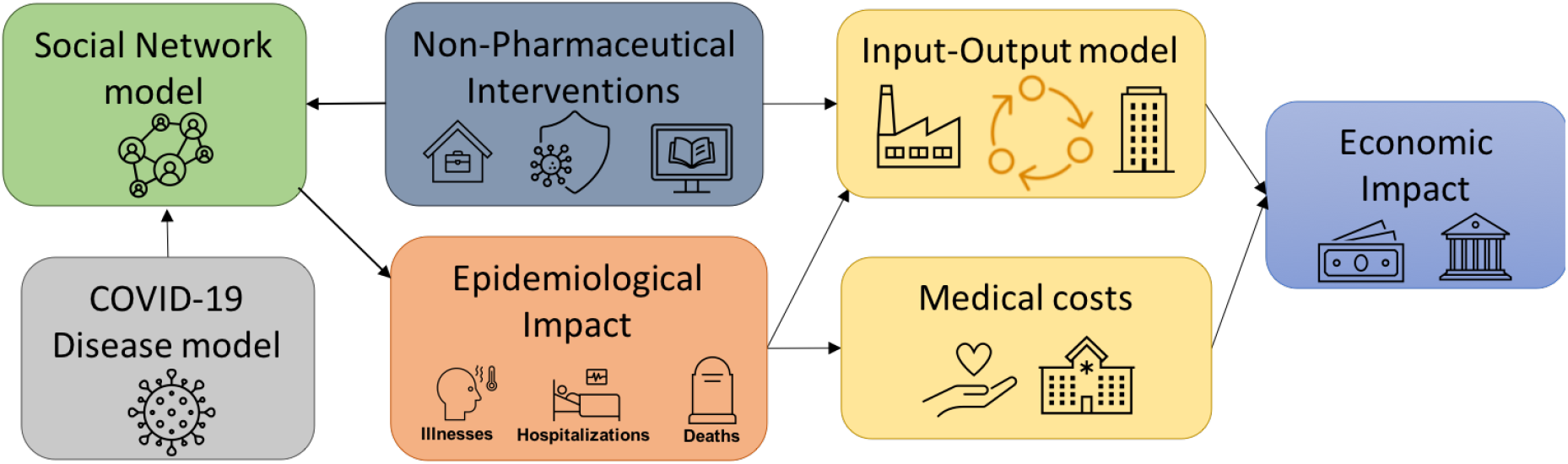
This figure shows the overall modeling framework, its various components and their linkages.

### 3.1 Social Contact Networks

We use a synthetic social contact network generated using the methodology provided in [5, 9, 24] and used in [2, 13–15, 19, 23, 28, 36] to study the spread of COVID-19. An agent based model is used to build an individual level social contact network based on the colocation of the individuals over the course of a day. These types of networks have been validated and used to study various infectious diseases, interventions, and their public health implications. For details on these studies and on the methodology to generate synthetic social contact networks, see [5, 6, 14, 22–24, 29, 34].

Each individual in the social network is endowed with a list of demographic attributes such as age, gender, income, occupation, family size, family income etc. consistent with the data provided by the US Census. A person’s occupation and the associated sector to which the occupation is linked, along with the health state (susceptible, infected or dead) of the person, are used to determine the sector level interruption in labor supply on any day that arises from sickness, mortality or stay-home order. This is the critical piece that joins the disease model with the economic model.

### 3.2 Disease Model

The disease model is the *best guess version* of “COVID-19 Pandemic Planning Scenarios” prepared by the US Centers for Disease Control and Prevention (CDC) SARS-CoV-2 Modeling Team [12]. It is an SEIR model where state transitions follow the parameters as defined in the document. The disease states and transition paths are shown in figure 2. The final disease state can be reached through multiple paths. The model is also age stratified for the following categories i.e. preschool (0-4 years), students (5-17) adults (18-49), older adults (50-64) and seniors (65+) and calibrated for each of the age groups separately. More details on the disease model are available in the Supplementary Information.

**Figure 2:**
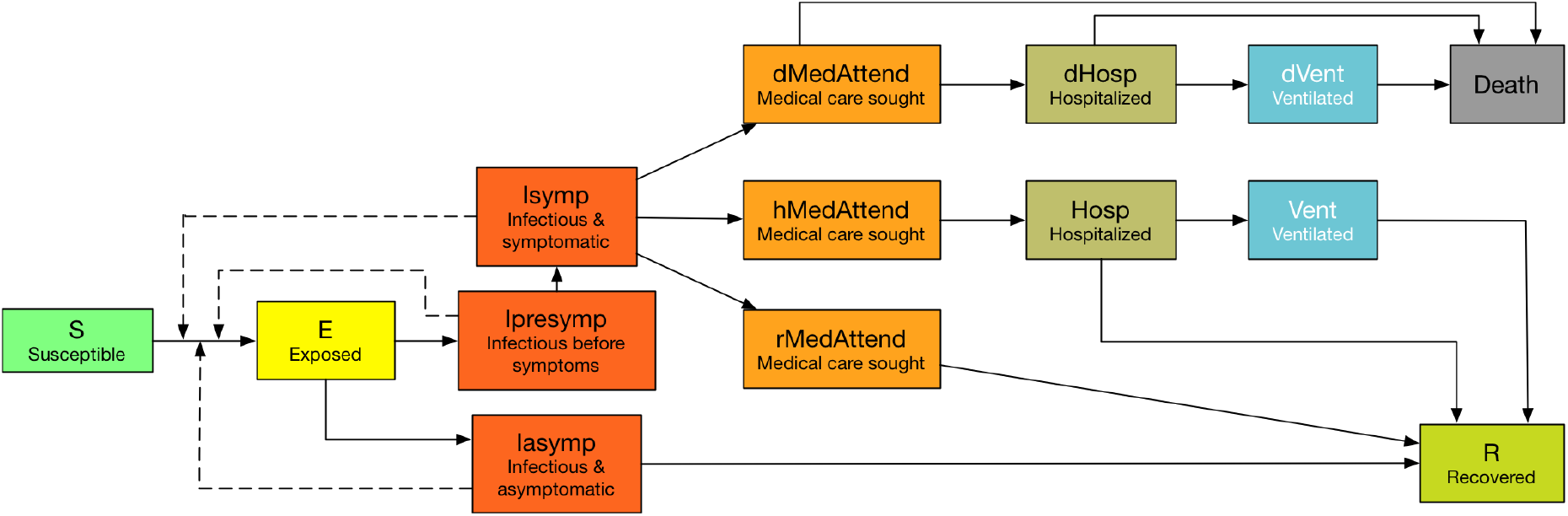
Disease states and transition paths in the COVID-19 disease model.

### 3.3 Non-Pharmaceutical Interventions

We apply a number of social distancing strategies to mitigate the spread of COVID-19 [15]. We assume there are no vaccines available and non-pharmaceutical interventions (NPI) are the only way to control the spread of COVID-19. We use the following NPI strategies: (i) **Voluntary home isolation (VHI)** – symptomatic people choose to stay at home (non-home type contacts are disabled) for 14 days. (ii) **School closure (SC)** – schools and colleges are closed (school type contacts are disabled). (iii) **Stay home (SH)** – a lockdown order directs people to “stay-home” (non-home type contacts are disabled).

School closure and stay-home interventions start on different days in different states as stated in [20, 39]. Once closed, schools are assumed to remain closed until end of August. The duration and compliance to social distancing measures vary across scenarios as shown in table 1.

**Table 1:** Terminology of counterfactual scenarios: VHI and SH refer to “voluntary home isolation” and “stay-home” order respectively. VHI and SH compliance rates can vary between 60% and 90% and the duration of SH order can be 30, 45 or 60 days. The first two numbers in the scenario name indicate compliance rate and the last one indicates the duration of the stay-home order.

| SH duration<br>(in days) | VHI and SH compliance rates |  |  |  |
| --- | --- | --- | --- | --- |
|  | 60% | 70% | 80% | 90% |
| 0 | None (unmitigated) |  |  |  |
| 30 | VHI_60_SH_60_30 | VHI_70_SH_70_30 | VHI_80_SH_80_30 | VHI_90_SH_90_30 |
| 45 | VHI_60_SH_60_45 | VHI_70_SH_70_45 | VHI_80_SH_80_45 | VHI_90_SH_90_45 |
| 60 | VHI_60_SH_60_60 | VHI_70_SH_70_60 | VHI_80_SH_80_60 | VHI_90_SH_90_60 |

Stay-home durations are set at 0, 30, 45 to 60 days. Compliance to SH and VHI are set at 60%, 70%, 80% and 90%. Table 1 lists all the scenarios including the unmitigated one. For each experimental cell, 25 simulation replicates are run and results are shown based on the average values across these runs. Table 2 shows the parameters used in the experiments for easy reference.

**Table 2:**
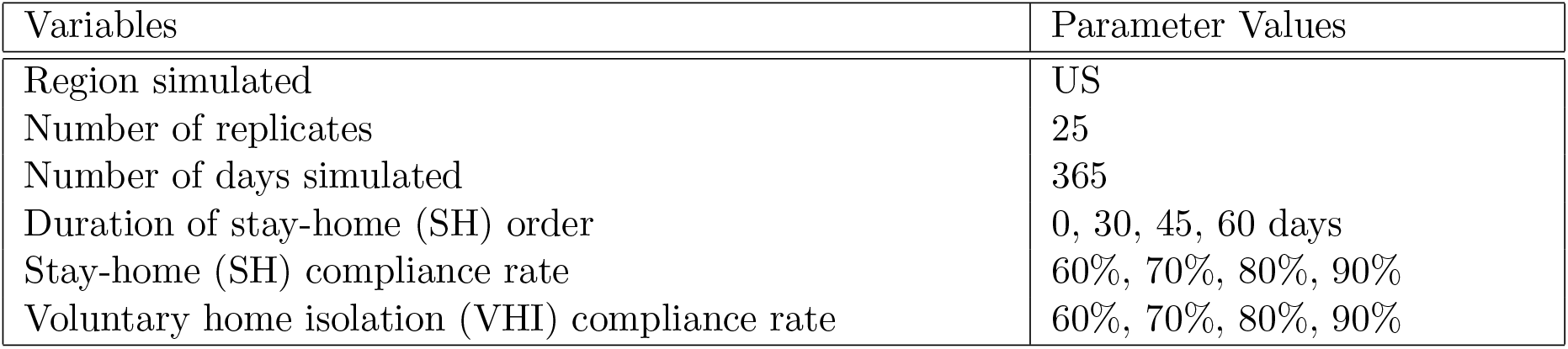
Variables used in the simulation experiments.

### 3.4 Medical Costs

Medical cost of treating COVID-19 patients under different health states are taken from [15, 33], which provide the average payment for treating pneumonia cases among “large employer health insurance” plans, and under different severity levels. See table 3. In the absence of COVID-19 treatment cost data, the pneumonia estimates have been used as a proxy. Note that each infected individual’s medical cost is counted only once. For example if a person is in ventilated state, after having gone through “medAttend” and “Hosp” state, costs are cumulative to the “vent” state [15]. To estimate the medical costs of COVID-19 for each scenario, we multiply the number of medically attended, hospitalized, and ventilated with the estimated treatment costs per person given in table 3. This is repeated for each replicate in the simulation and the average estimates are reported. Note that an earlier paper focuses entirely on the medical costs [15] and provides more details on medical costs to the interested reader.

**Table 3:**
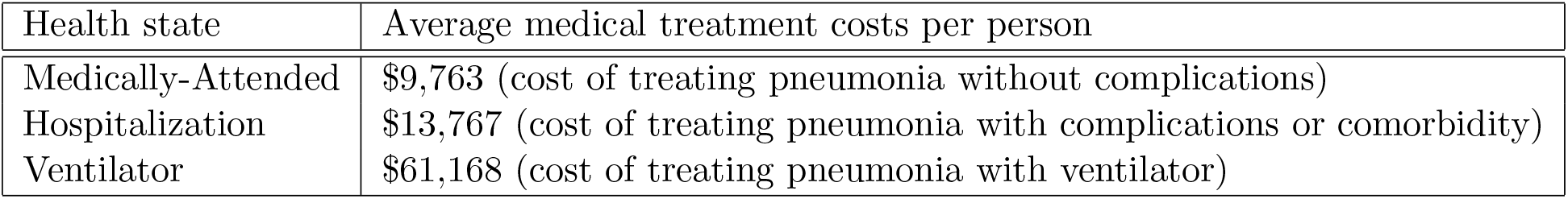
Average cost of medical care under different health states [33].

### 3.5 Economic Sectors and their Interdependencies

We use the summary level input-output (I-O) tables for 2018 downloaded from the US Bureau of Economic Analysis (BEA) [11], which quantify how industries depend on each other and interact with each other, to capture the cascading effect of labor supply shock across industries. The entire economy is divided into 71 industries; the I-O data reflects the structure of the US economy and the relative importance of each industry with respect to all other industries. We follow the NAICS (North American Industry Classification System) codes to aggregate the I-O data to sector level (21 sectors). Figure 3 shows the interdependencies between the 21 sectors. The flows from sectors on the left of figure 3 are input to sectors on the right of the figure.

**Figure 3:**
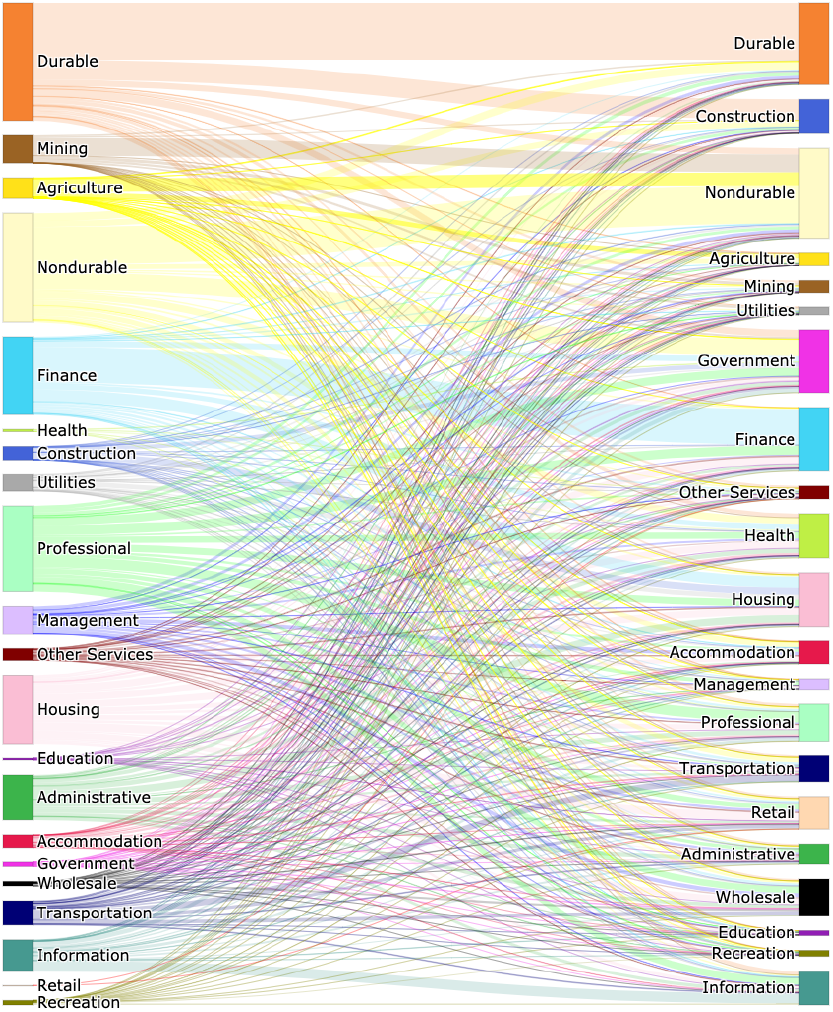
This figure shows interdependencies between sectors as given by the US Bureau of Economic Analysis. The left sector flows are input to right sectors.

### 3.6 Data on Telework by Sector

During the stay-home order, some individuals are able to work from home. However, the ability to work from home (WFH) and the productivity of WFH workers vary by the type of sector the individuals are employed in. Authors in [8, 18] estimate the number of jobs that can be done from home in the US. Work in [18] combines the feasibility of working from home by occupation, with occupational employment counts, and determines that 37% of all jobs in the US can be done from home.

Although this is not uniform across all sectors and cities; sectors like computing, education, legal and financial can be largely operational from home but construction, farming and hospitality cannot be. [18] provides the fraction of jobs that can be done from home by NAICS (North American Industry Classification System) and by SOC (Standard Occupational Classification) occupation. We use this fraction for each sector to determine the fraction of labor that can work from home. In addition, [18] provides the fraction of teleworkable wages for each sector. Together, these fractions determine the level of productivity that can be maintained during a lockdown by the healthy workforce in each sector. The health of each individual is tracked by the disease model given in section 3.2.

### 3.7 Input-Output Model

We use the Dynamic Inoperability Input-Output Model (DIIM) stated in [27,32] to study the effect of labor supply shock arising from the morbidity and mortality caused by COVID-19, as well as the enforcement of the stay-home order, on national productivity. The DIIM model uses the classic input-output (I-O) economic analysis of Leontief (1935) [26] to account for the interdependencies between sectors. Additionally, it allows modeling of resiliency parameters within the I-O model to signify sector wise recovery rates. We use the DIIM model to estimate the direct effect of drop in labor supply to each sector due to sickness, deaths and lockdown, as well as the indirect effect to sectors that arise due to interdependencies between sectors.

Depending upon the scenario considered in the simulation, appropriate interventions are applied to the social network. The interventions result in removal of edges on a temporal basis in the social contact network. For example, a stay-home order results in removal of all non-home edges of the compliant individuals for the duration of the order. The COVID-19 disease model is seeded and run on this time-varying social contact network over a period of one year. Everyone in the population is assumed to be susceptible at the beginning of the simulations except the seed nodes or the index cases, which are assumed to be infected. As the disease spreads through the network, the simulations generate a time series of daily infections. The infected individuals are further divided into medically attended, hospitalized, ventilated and dead, based on the probabilities assumed in the disease model.

To calculate the labor supply shock to each sector and its impact on productivity, we estimate (i) the number of infected and dead each day in each sector (using occupation and NAICS codes) and calculate the fraction of labor that is unable to work; (ii) the healthy individuals who comply with the stay-home order and do not go to work, and also cannot work from home given their occupation-type, as determined by telecommuting data for each sector [18]; and (iii) healthy individuals who can work from home but their productivity is reduced as suggested in the teleworkable wages for each sector in [18].

## 4 Results and Discussion

We calculate the economic losses under the unmitigated scenario and the mitigation scenarios. Mitigation efforts help control the spread of the disease and hence reduce the total number of infections but they also increase the economic losses due to social distancing measures like the stay-home order. The compliance to NPIs and the length of the NPIs determine the extent of the loss, which can be weighed against the benefits measured in terms of reduced number of infections and deaths.

### 4.1 Economic losses due to inoperability and NPIs

Figure 4 shows the economic losses due to the inoperability of sectors under different NPIs and the infections caused by the pandemic. The left subfigure does not include the economic burden imposed by the treatment and medical services given to the infected individuals.

**Figure 4:**
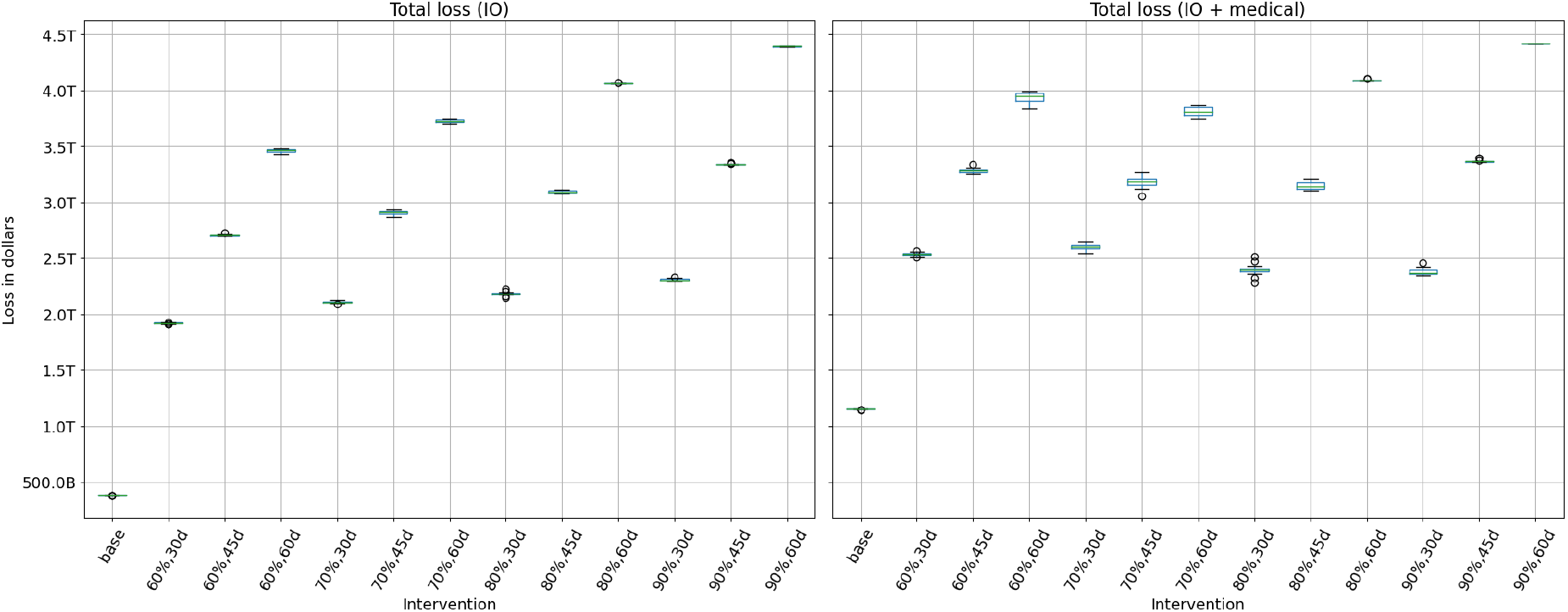
The left subfigure shows I-O economic losses (without medical costs) due to NPI measures and infections for each of the scenarios. The percentages show compliance to NPIs and “d” is for the duration of the Stay-home order. These losses arise from the drop in labor supply to sectors, caused by lockdown, illness and mortality, and from interdependencies between sectors. It does not include the economic burden imposed by the treatment and medical services provided to the infected individuals, whereas the right subfigure includes this medical burden.

Lockdown and other social distancing measures reduce the labor supply to sectors but these measures do not uniformly affect each sector’s output. Depending upon how labor-intensive a sector is, how many jobs can be done from home, and how much value each job generates in a sector, the lockdown has a differential impact on each sector. For example, Education, Professional services, and Management sectors are teleworkable at 80% or higher levels whereas Accommodation (includes hospitality and food services) is at 3% and Agriculture is at 7%.

Inability to work from home in Construction and Agriculture sectors should imply more losses in these sectors. However we find that the losses are higher in Education, Professional services and Management sectors because jobs in these sectors pay more on average than the jobs in Construction and Agriculture sectors. Hence even a 20% loss in work in the former sectors can result in a higher total loss in value compared to a 90% loss in work in the latter sectors.

Overall economic loss from inoperability also depend on the level of dependency each sector has on others. Agriculture and Construction sectors also have a higher level of dependency on other sectors compared to Education, Professional and Management sectors as shown in figure 3. The lack of self-reliance increases the potential for losses caused by the cascading effect of other sectors. The results in left subfigure 4 show that as the duration of SH order increases, the economic losses increase for a given compliance rate. This is because a longer SH order implies that healthy individuals are not able to work. A longer SH order also reduces the number of infections and deaths and hence improves labor supply and productivity. There is less absenteeism due to sickness and death, and less cascading effect on other sectors. The overall drop in productivity from a longer SH order shows that the gain in productivity from fewer infections and deaths is less than the loss from a longer shutdown. However SH order saves tens of thousands of lives and millions of infections as described in Section 4.4.

In the unmitigated base case, the economic loss is low but the loss due to morbidity and mortality is high. The healthy individuals are assumed to be working in the unmitigated scenario since no NPIs are in effect. The drop in productivity is caused only by the drop in labor supply due to illness and deaths since there is no lockdown in place. However, in the unmitigated case, more than 117,000 lives are lost and over 116 million infections occur.

### 4.2 Economic losses due to inoperability, NPIs and medical treatments

In figure 4, the right subfigure shows the losses that are included in the left subfigure 4 plus the economic burden caused by the medical treatment of the ill. Note that the total loss in the unmitigated case without medical costs is $0.38 trillion in left subfigure 4 whereas with medical costs, this loss increases to $1.15 trillion as shown in the right subfigure 4. The extra $0.8 trillion is solely due to the medical costs of treating infections in the unmitigated case. Note that for a given compliance level, a longer SH always results in a higher loss. However for a given SH duration, a higher compliance may result in a lower or higher loss. This would depend upon the relative gain from reduced infections versus loss from SH of healthy individuals.

For example, in right subfigure 4 when the SH duration is set at 60 days, increasing compliance from 60% to 70% decreases the economic loss but increasing compliance from 70% to 80% increases the economic loss. This is because compliance has a non-linear effect on losses. At low levels of compliance, the marginal effect of a small increase in compliance is high because it helps get the pandemic under control which implies less absenteeism due to illness and lower medical costs. An increase in compliance from 70% to 80% does not have the same incremental effect on infections because 70% compliance is already effective, but has a large effect on the inoperability of sectors because a larger critical mass of workers are staying home.

### 4.3 Trade-offs Between Compliance and Duration of Lockdown

Both subfigures in figure 4 show that there are tradeoffs between compliance and the length of the SH order. Low compliance can be compensated by a longer SH order and a shorter SH order can be combined with a higher compliance level to reach the same level of total loss. For example, in left subfigure 4, a 60% compliance rate combined with a 60 days of SH results in similar total loss as a 90% compliance rate combined with a 45 days of SH.

The best outcome is reached when the lockdown is for 30 days and the compliance at least 80%, as shown in right subfigure 4. It is clear that a lengthy SH order is harmful to the economy so a short SH order combined with a high level of compliance is ideal. Note that these tradeoffs and losses do not include the long term effect of deaths, i.e. the permanent loss in productivity, and only consider loss in labor supply for the duration of the simulation. The number of deaths depend on the duration and compliance to NPIs and are an important metric in measuring the outcomes. Later plots show the number of infections and deaths averted under each scenario.

### 4.4 Infections and Deaths Averted Versus Economic Loss

Figures 5 and 6 show the trade-off between the number of infections-averted and economic losses, as well as the number of deaths-averted vs. economic losses respectively, under each of the intervention scenarios. Several important observations can be made from these plots: (i) The base case, where no NPIs are in place has the least loss but results in over 100 million infections and over 100,000 deaths. (ii) The trend for both morbidity and mortality is the same under different scenarios. (iii) Losses rise with longer durations of SH order. (iv) A SH order of 45 days results in same economic loss whether the compliance is at 70% or 80%. However the numbers of infections and deaths averted are much higher at 80% compliance. (v) Similarly, once 90% compliance is reached, an increase in SH duration from 45 to 60 days does not reduce infections and deaths but adds more than one trillion in economic losses. (vi) A longer lockdown can compensate for the lack of compliance and a higher compliance can reduce the duration of the lockdown in order to achieve similar number of infections and deaths. However these trade-offs are non-linear and these kinds of analytics are needed to inform public health policy.

**Figure 5:**
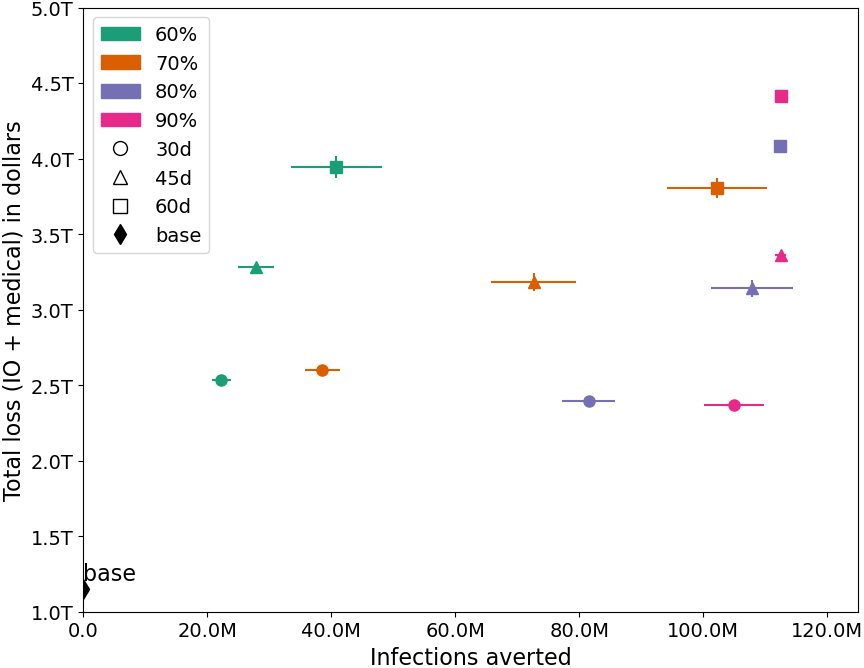
Trade-off between the number of infections averted and economic losses under each scenario. The vertical and horizontal bars show the inter-quartile range.

**Figure 6:**
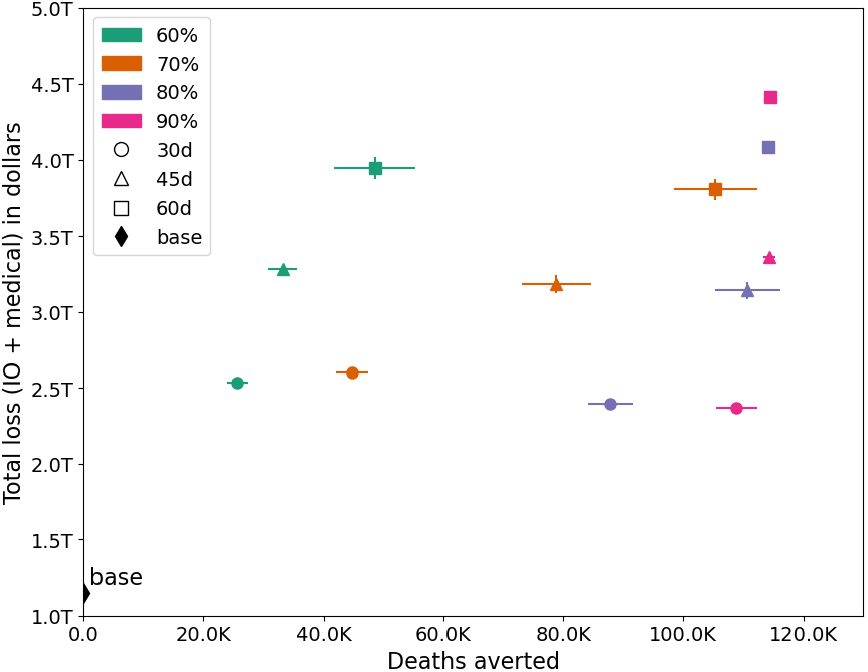
Trade-off between the number of deaths averted and economic losses under each scenario. The vertical and horizontal bars show the inter-quartile range.

### 4.5 Sector Level Economic Losses

We calculate sector level losses to understand how each sector would be impacted under different intervention scenarios. Figure 7 shows the daily loss across all sectors for each of the scenarios, including the unmitigated one. The percentages show compliance to VHI and SH and “d” reflects the duration of SH order. On the top the curves are clustered by the duration of SH order. The longer the SH order, the wider the top is; reflecting a more sustained loss at peak level during the lockdown period. Note that a second peak occurs only in scenarios where the compliance is low or compliance and duration both are low. As expected, the economic loss is higher in all intervention scenarios compared to the unmitigated scenario, since NPIs are in effect which keep healthy people from going to work. However, as shown in figures 5 and 6, these NPIs are able to avert over 100 million infections and over 100,000 deaths.

**Figure 7:**
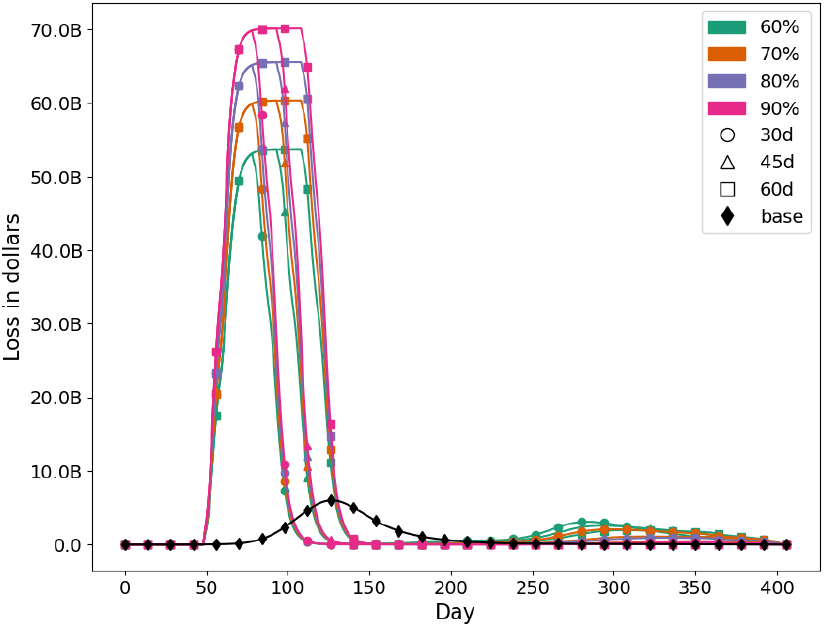
**Daily I-O loss** across all sectors for each scenario. The unmitigated base case is shown by the black curve where no NPIs are in place. Percentages are compliance to NPIs and “d” is the duration of SH order.

Figure 8 shows total loss for each sector and for each scenario, across time. The unmitigated case, marked in black, shows the least amount of economic loss since there are no NPIs in place. For comparative analysis, we select a medium level scenario, *V HI_*70_ *SH_*70_45, and discuss in more detail. This is highlighted and marked in orange. In most of the cases, the sectors that are biggest losers are Government, Durables, Health and Non-durables.

**Figure 8:**
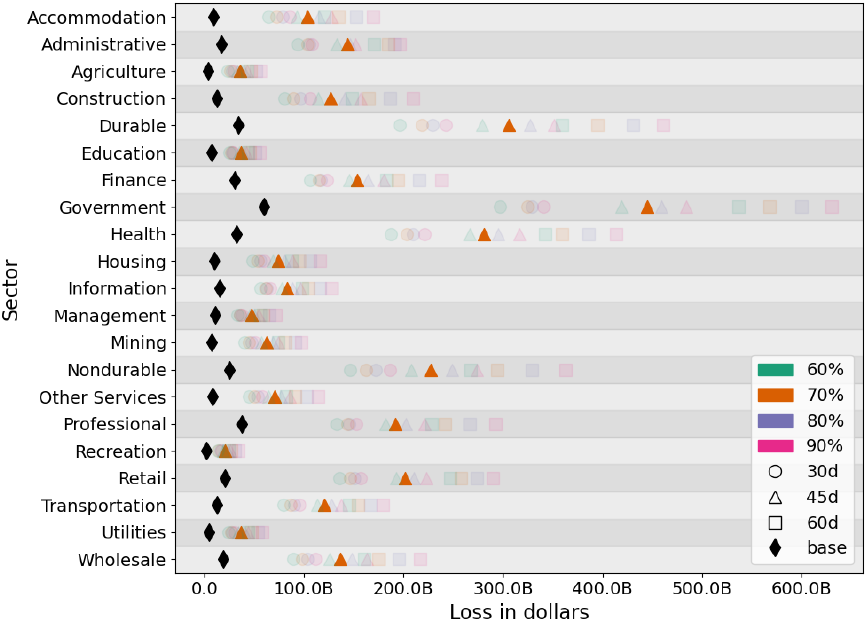
Sector level **total loss** for each scenario. The black and orange markers are highlighted to show the unmitigated and mitigated case *V HI_*70_ *SH_*70_45 respectively. Percentages are compliance to NPIs and “d” is the duration of SH order.

Figure 9 shows a detailed comparison of sectoral loss for the unmitigated and a mitigated scenario over time. These do not include any medical costs. The left subfigure shows that without any mitigation, the highest losses occur in Government, Professional, Durables, Health and Finance sectors. Note that these losses are caused by loss in labor force due to sickness and deaths. There are no NPIs in effect in the unmitigated case. Even though inoperability is higher in sectors like Agriculture, Construction and Accommodation which tend to be more labor intensive, the value generated by the same proportional loss in labor is higher in Government, Professional, Durables, Health and Finance sectors due to their higher per capita productivity.

**Figure 9:**
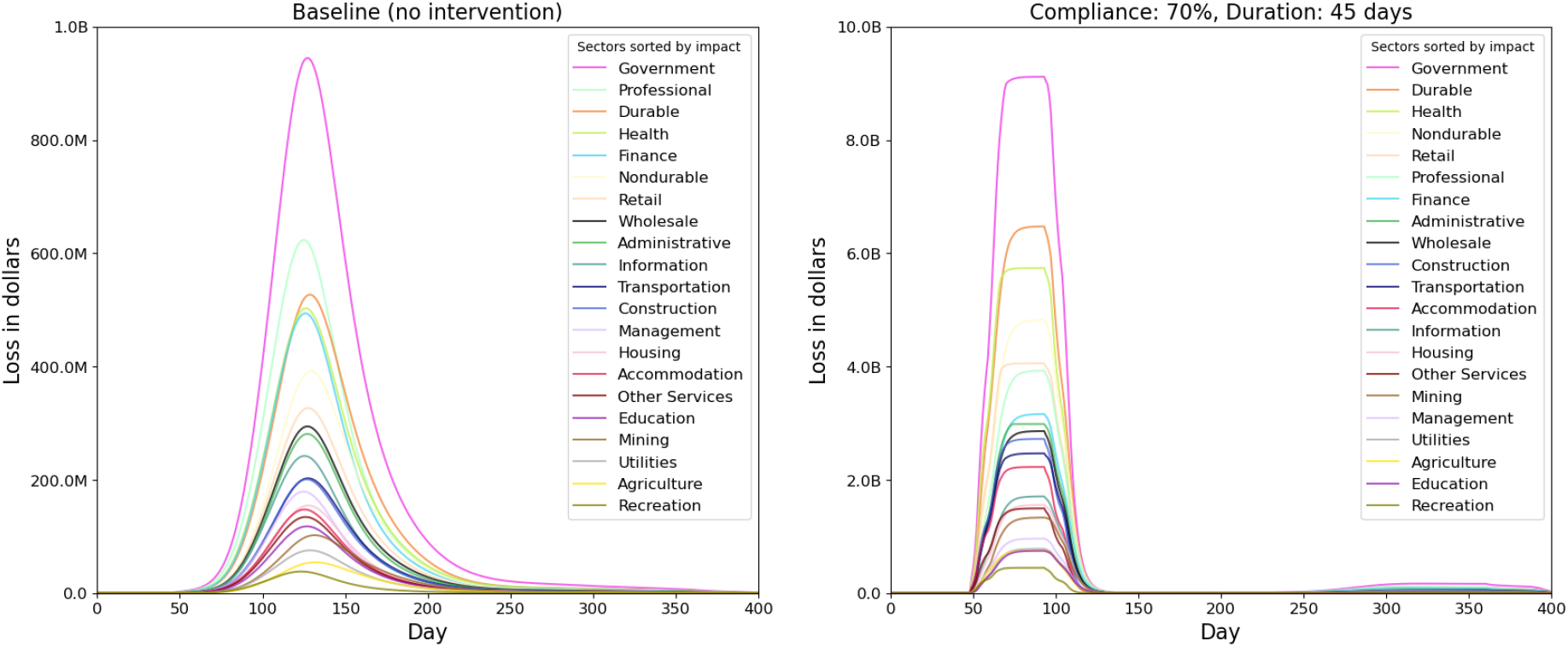
Sector level daily losses caused by the inoperability of each sector and its cascading impact on other sectors due to interdependencies between sectors. The left subfigure shows losses for the unmitigated case. The right subfigure shows losses for the mitigated scenario VHI_70_SH_70_45. The ordering of sectors in the legend is ranked by the height of the curve. Note that the scale of loss (y-axis) in the right subfigure is ten times of the left subfigure.

#### 4.5.1 Detailed Analysis of an Intervention Scenario

Here we provide a detailed sector level analysis of one of the 12 mitigation scenarios. We pick *V HI* 70 *SH* 70 45 as an example case since it represents a mid-level scenario. The right subfigure in figure 9 shows daily losses in each of the sectors under this scenario and relative rankings of sectors when NPIs are in effect. The top 5 sectors in terms of biggest economic losses are Government, Durables, Non-durables, Health and Retail.

Even though the inoperability in these sectors is not that high due to NPIs, these sectors have higher wages and represent higher values compared to sectors which are more labor intensive. Top 5 sectors that have the highest inoperability due to labor supply shock from mitigation are Accommodation, Retail, Agriculture, Transportation and Construction but their losses are relatively low because of the low wages in these sectors. Other major factors that affect the losses in each sector are the extent to which the employment and wages are teleworkable. For example, in the unmitigated case, the worst performers include Finance because even a small shock to labor supply in this sector causes a big loss in value compared to a similar shock to sectors like Agriculture; but in the mitigated case, Finance sector performs relatively better because 76% of its jobs and 85% of the wages in Finance are teleworkable, whereas in Agriculture it is only 7% and 13% respectively.

Even in the mitigated case the Government sector has the highest loss, partly because it is also the largest sector in the economy but partly because it has a very high dependency on Durables, Non-durables and Professional which themselves are hit hard. Additionally in the Government sector, only 41% of the jobs and 46% of the wages are teleworkable,

### 4.6 Best Mitigation Scenario

The best mitigation scenario in terms of lives saved and infections averted is when the compliance is at 90% and SH duration is 45 days. See figures **??** and 6. This scenario results in a total economic loss of about $3.4 trillion dollars. However, it also saves more than 110,000 lives and 115 million infections compared to the unmitigated case. Assuming US federal government’s estimate of value of life which is $10 million per person [3, 30], lowering the number of deaths would save $1.1 trillion and lowering number of infections would save medical costs equivalent to $0.8 trillion, resulting in a gain of about $1.9 trillion from the mitigation efforts and a net economic loss of $1.5 trillion. This kind of simulation based analysis can help prioritize epidemiological and economic goals, understand their trade-offs, and guide public health policy.

## 5 Limitations

This study does not consider the demand side shock to the economy that results in drop in demand for goods and services due to lower employment, lost wages, and uncertain economic conditions. Unlike the general equilibrium model where demand and supply shock result in price adjustment, the input-output model does not capture the price dynamics that arise from changes in demand and supply. The treatment costs are average costs for treating pneumonia patients as available from [33] which do not vary by age, but only by severity of the case and these are used as proxies for COVID-19 costs.

## 6 Summary and Conclusions

This study estimates the epidemiological and economic impact of several counterfactual intervention scenarios to contain the spread of COVID-19. Results show that any intervention involving a stayhome order will result in significant economic losses. However, the epidemiological impact of these interventions is dramatic. We find that interventions scenarios involving 45 days of SH order and a high compliance to NPIs can save more than 110,000 lives and 115 million infections compared to the unmitigated case.

We perform a sector level impact analysis and find that losses depend on the level of labor supply shock, the ability of employees to work from home, the productivity of workers who work from home and the dependency on other sectors. The sectors that are more labor intensive such as Agriculture and Construction are not the worst performers because the per capita value generated is lower in these sectors compared to sectors like Government and Health.

Our results also show trade-offs between the economic losses and the number of deaths and infections averted. A longer lockdown and/or a high compliance to NPIs result in higher economic losses but save lives and reduce the number of COVID-19 infections. There is also a trade-off between duration of the lockdown and the rate of compliance to NPIs. If people are non-compliant to NPIs, public health policy-makers can increase the duration of the lockdown to get the same level of results in terms of infections and deaths averted.

## Data Availability

All the output data reported in the paper is available upon request, but restrictions apply on the commercially available data used in the construction of the social contact network and hence the availability of the social network data itself.

## Code Availability

Code developed to analyze the results and support the findings in this paper is available upon request, from the corresponding author.

## Acknowledgements

This work was partially supported by National Institutes of Health (NIH) Grant R01GM109718, NSF BIG DATA Grant IIS-1633028, NSF DIBBS Grant ACI-1443054, NSF Grant No.: OAC-1916805, NSF Expeditions in Computing Grant CCF-1918656, CCF-1917819, US Centers for Disease Control and Prevention 75D30119C05935, DTRA subcontract/ARA S-D00189-15-TO-01-UVA, and collaborative seed grants from the UVA Global Infectious Disease Institute. The content is solely the responsibility of the authors and does not necessarily represent the official views of the sponsoring agencies.

## Contributions

JC, SH, HM, SE, SV, JS and BL built the model and the software. AV, JC, JS, SV, WY and AM processed and analyzed the data. AV, JC, AM, MM and CB conceived the project. All authors helped write, edit and review the paper.

## Additional Information

The authors have no competing interests as defined by Nature Research, or other interests that might be perceived to influence the results and/or discussion reported in this paper.

## Supplementary Information

### The Disease Model Parameters

The CDC disease model in Figure 1 shows the state transitions which include transmissions and progressions. The former occurs when an individual in *Susceptible* state comes in contact with an individual in one of *Presymptomatic, Symptomatic*, or *Asymptomatic* states. The latter occurs when an individual has been in that state for a certain amount of time (called dwell time); the transitions are probabilistic. The dwell time distributions and the transition probability distributions are age dependent and are specified in the table.

| Progression | Attribute | Age group |  |  |  |  |
| --- | --- | --- | --- | --- | --- | --- |
|  |  | 0-4 | 5-17 | 18-49 | 50-64 | 65+ |
| Exposed - Asympt | prob | 0.35 |  |  |  |  |
| Exposed - Asympt | dt-mean | 5 |  |  |  |  |
| Exposed - Asympt | dt-std dev | 1 |  |  |  |  |
| Asympt - Recovered | prob | 1 |  |  |  |  |
| Asympt - Recovered | dt-mean | 5 |  |  |  |  |
| Asympt - Recovered | dt-std dev | 1 |  |  |  |  |
| Exposed - Presympt | prob | 0.65 |  |  |  |  |
| Exposed - Presympt | dt-fixed | 1 |  |  |  |  |
| Presympt - Sympt | prob | 0.65 |  |  |  |  |
| Presympt - Sympt | dt-fixed | 1 |  |  |  |  |
| Sympt - Attd | prob | 0.9594 | 0.9894 | 0.9594 | 0.912 | 0.788 |
| Sympt - Attd | dt-discrete | 1:0.175, 2:0.175, 3:0.1, 4:0.1, 5:0.1, 6:0.1, 7:0.1, 8:0.05, 9:0.05, 10:0.05 |  |  |  |  |
| Attd - Recovered | prob | 1 |  |  |  |  |
| Attd - Recovered | dt-mean | 5 |  |  |  |  |
| Attd - Recovered | dt-std dev | 1 |  |  |  |  |
| Sympt - Attd(D) | prob | 0.0006 | 0.0006 | 0.0006 | 0.003 | 0.017 |
| Sympt - Attd(D) | dt-fixed | 2 |  |  |  |  |
| Attd(D) - Hosp(D) | prob | 0.95 |  |  |  |  |
| Attd(D) - Hosp(D) | dt-fixed | 2 |  |  |  |  |
| Hosp(D) - Vent(D) | prob | 0.06 | 0.06 | 0.06 | 0.15 | 0.225 |
| Hosp(D) - Vent(D) | dt-fixed | 2 |  |  |  |  |
| Vent(D) - Death | prob | 1 |  |  |  |  |
| Vent(D) - Death | dt-fixed | 4 |  |  |  |  |
| Hosp(D) - Death | prob | 0.94 | 0.94 | 0.94 | 0.85 | 0.775 |
| Hosp(D) - Death | dt-fixed | 6 |  |  |  |  |
| Attd(D) - Death | prob | 0.05 |  |  |  |  |
| Attd(D) - Death | dt-fixed | 8 |  |  |  |  |
| Sympt - Attd(H) | prob | 0.04 | 0.01 | 0.04 | 0.085 | 0.195 |
| Sympt - Attd(H) | dt-fixed | 1 |  |  |  |  |
| Attd(H) - Hosp | prob | 1 |  |  |  |  |
| Attd(H) - Hosp | dt-mean | 5 | 5 | 5 | 5.3 | 4.2 |
| Attd(H) - Hosp | dt-std dev | 4.6 | 4.6 | 4.6 | 5.2 | 5.2 |
| Hosp - Recovered | prob, | 0.2 |  |  |  |  |
| Hosp - Recovered | dt-mean | 3.1 | 3.1 | 3.1 | 7.8 | 6.5 |
| Hosp - Recovered | dt-std dev | 3.7 | 3.7 | 3.7 | 6.3 | 4.9 |
| Hosp - Vent | prob | 0.06 | 0.06 | 0.06 | 0.15 | 0.225 |
| Hosp - Vent | dt-mean | 1 |  |  |  |  |
| Hosp - Vent | dt-std dev | 0.2 |  |  |  |  |
| Vent - Recovered | prob | 1 |  |  |  |  |
| Vent - Recovered | dt-mean | 2.1 | 2.1 | 2.1 | 6.8 | 5.5 |
| Vent - Recovered | dt-std dev | 3.7 | 3.7 | 3.7 | 6.3 | 4.9 |

Age dependent dwell-time distributions and transition probability distributions. Here “dt” refers to duration, e.g. dt-mean is mean duration and dt-discrete is discrete distribution duration. “Attd” is medically attended; “Hosp” is hospitalized; “Vent” is ventilated; “Sympt” is symptomatic and “Asympt” is asymptomatic.

## References

[1] D. Acemoglu, V. Chernozhukov, I. Werning, and M. D. Whinston. Optimal targeted lockdowns in a multi-group sir model. NBER Working Paper, 27102, 2020.

[2] A. Adiga, S. Venkatramanan, J. Schlitt, and et al. Evaluating the impact of international airline suspensions on the early global spread of covid-19. Technical report, medRxiv, 2020. https://www.medrxiv.org/content/10.1101/2020.02.20.20025882v2.full.pdf.

[3] F. E. Alvarez, D. Argente, and F. Lippi. A simple planning problem for covid-19 lockdown. Technical report, National Bureau of Economic Research, 2020.

[4] S. R. Baker, N. Bloom, S. J. Davis, and S. J. Terry. Covid-induced economic uncertainty. Technical report, National Bureau of Economic Research, 2020.

[5] C. Barrett, R. Beckman, M. Khan, V. S. Anil Kumar, M. Marathe, P. Stretz, T. Dutta, and B. Lewis.Generation and analysis of large synthetic social contact networks. In Winter Simulation Conference, pages 1003–1014. Winter Simulation Conference, 2009.

[6] C. Barrett, S. Eubank, and M. Marathe. An interaction-based approach to computational epidemiology. In AAAI, pages 1590–1593, 2008.

[7] R. J. Barro, J.F. Ursuá, and J. Weng. The coronavirus and the great influenza pandemic: Lessons from the “spanish flu” for the coronavirus’s potential effects on mortality and economic activity. Technical report, National Bureau of Economic Research, 2020.

[8] A. Bick, A. Blandin, and K. Mertens. Work from home after the covid-19 outbreak. 2020.

[9] K. Bisset and M. Marathe. A cyber-environment to support pandemic planning and response. DOE SciDAC Magazine, pages 36–47, 2009.

[10] M. Brahmbhatt and A. Dutta. On SARS type economic effects during infectious disease outbreaks. The World Bank, 2008.

[11] Bureau of Economic Analysis. Input-output accounts data. https://www.bea.gov/industry/input-output-accounts-data. Accessed on April 30, 2020.

[12] Centers for Disease Control and Prevention. Covid-19 pandemic planning scenarios. https://www.cdc.gov/coronavirus/2019-ncov/hcp/planning-scenarios-h.pdf, 2020. [Online, accessed July 8, 2020].

[13] J. Chen, S. Levin, S. Eubank, H. Mortveit, S. Venkatramanan, A. Vullikanti, and M. Marathe. Networked epidemiology for covid-19. SIAM News, 53(5):3,7, june 2020.

[14] J. Chen, A. Marathe, and M. Marathe. Feedback between behavioral adaptations and disease dynamics. Scientific reports, 8(1):1–15, 2018.

[15] J. Chen, A. Vullikanti, S. Hoops, H. Mortveit, B. Lewis, S. Venkatramanan, W. You, S. Eubank, M. Marathe, C. Barrett, and A. Marathe. Medical costs of keeping the us economy open during covid-19. Scientific reports, 10(1):1–10, 2020.

[16] O. Coibion, Y. Gorodnichenko, and M. Weber. Labor markets during the covid-19 crisis: A preliminary view. Technical report, National Bureau of Economic Research, 2020.

[17] S. Correia, S. Luck, and E. Verner. Pandemics depress the economy, public health interventions do not: Evidence from the 1918 flu. Public Health Interventions Do Not: Evidence from the, 1918.

[18] J. I. Dingel and B. Neiman. How many jobs can be done at home? Journal of Public Economics, 2020.

[19] N. Dorratoltaj, A. Marathe, B. L. Lewis, S. Swarup, S. G. Eubank, and K. M. Abbas. Epidemiological and economic impact of pandemic influenza in chicago: Priorities for vaccine interventions. PLoS computational biology, 13(6):e1005521, 2017.

[20] Education Week. Map: Coronavirus and school closures. https://www.edweek.org/ew/section/multimedia/map-coronavirus-and-school-closures.html, 2020. [Online, accessed June 30, 2020].

[21] M. S. Eichenbaum, S. Rebelo, and M. Trabandt. The macroeconomics of epidemics. Working Paper 26882, National Bureau of Economic Research, March 2020.

[22] S. Eubank, C. L. Barrett, R. Beckman, K. Bisset, L. Durbeck, C. Kuhlman, B. Lewis, A. Marathe, M. Marathe, and P. Stretz. Detail in network models of epidemiology: Are we there yet? Journal of Biological Dynamics, 4:446–455, 2010. PubMed PMID: 20953340; PMCID: PMC2953274.

[23] S. Eubank, H. Guclu, V. S. Anil Kumar, M. Marathe, A. Srinivasan, Z. Toroczkai, and N. Wang. Modelling disease outbreaks in realistic urban social networks. Nature, 429:180–184, 2004.

[24] M. E. Halloran, N. M. Ferguson, S. Eubank, I. M. Longini, D. A. T. Cummings, B. Lewis, S. Xu, C. Fraser, A. Vullikanti, T. C. Germann, D. Wagener, R. Beckman, K. Kadau, C. Barrett, C. A. Macken, D. S. Burke, and P. Cooley. Modeling targeted layered containment of an influenza pandemic in the United States. In Proceedings of the National Academy of Sciences (PNAS), pages 4639–4644, March 10 2008. PMCID:PMC2290797.

[25] C. J. Jones, T. Philippon, and V. Venkateswaran. Optimal mitigation policies in a pandemic: Social distancing and working from home. Technical report, National Bureau of Economic Research, 2020.

[26] W. W. Leontief. Quantitative input and output relations in the economic systems of the united states. The review of economic statistics, pages 105–125, 1936.

[27] C. Lian and Y. Y. Haimes. Managing the risk of terrorism to interdependent infrastructure systems through the dynamic inoperability input–output model. Systems Engineering, 9(3):241–258, 2006.

[28] D. Machi, P. Bhattacharya, S. Hoops, J. Chen, H. Mortveit, S. Venkatramanan, B. Lewis, M. Wilson, A. Fadikar, T. Maiden, C. Barrett, and M. Marathe. Scalable epidemiological workflows to support covid-19 planning and response. Technical Report SC-TR-2020, Network Systems Science and Advanced Computing Division, BII, University of Virginia, 2020. https://www.dropbox.com/s/2uk2wnbte95mk5y/main.v87.7020f2c.pdf?dl=0.

[29] A. Marathe, B. Lewis, C. Barrett, J. Chen, M. Marathe, S. Eubank, and Y. Ma. Comparing effectiveness of top-down and bottom-up strategies in containing influenza. PLoS ONE, 6:e25149, 9 2011. PMCID: PMC3178616.

[30] D. Merrill. No one values your life more than the federal government. Bloomberg: https://www.bloomberg.com/graphics/2017-value-of-life, 2017.

[31] New York Times. Covid in the u.s.: Latest map and case count. https://www.nytimes.com/interactive/2020/us/coronavirus-us-cases.html, 2020. x[Online, accessed November 24, 2020].

[32] M. J. Orsi and J. R. Santos. Probabilistic modeling of workforce-based disruptions and input– output analysis of interdependent ripple effects. Economic Systems Research, 22(1):3–18, 2010.

[33] M. Rae, G. Claxton, N. Kurani, D. McDermott, and C. Cox. Potential costs of coronavirus treatment for people with employer coverage. Peterson Center on Healthcare and Kaiser Family Foundation, 13, 2020. https://www.healthsystemtracker.org/brief/potential-costs-of-coronavirus-treatment-for-people-with-employer-coverage/.

[34] C. Rivers, E. Lofgren, M. Marathe, S. Eubank, and B. Lewis. Modeling the impact of interventions on an epidemic of Ebola in Sierra Leone and Liberia. PLoS Currents, 2014. PMCID: PMC4399521.

[35] J. Santos. Perspectives on the effects of pandemic mitigation and suppression measures on interdependent economic sectors. Sustainable Production and Consumption, pages 249–255, 2020.

[36] M. Singh, P. Sarkhel, G. J. Kang, A. Marathe, K. Boyle, P. Murray-Tuite, K. M. Abbas, and S. Swarup. Impact of demographic disparities in social distancing and vaccination on influenza epidemics in urban and rural regions of the united states. BMC infectious diseases, 19(1):221, 2019.

[37] R. D. Smith, M. R. Keogh-Brown, T. Barnett, and J. Tait. The economy-wide impact of pandemic influenza on the uk: a computable general equilibrium modelling experiment. Bmj, 339:b4571, 2009.

[38] A. A. Toda. Susceptible-infected-recovered (sir) dynamics of covid-19 and economic impact. arXiv preprint arXiv:2003.11221, 2020.

[39] Wikipedia contributors. U.S. state and local government response to the covid-19 pandemic. https://en.wikipedia.org/wiki/U.S._state_and_local_government_response_to_the_COVID-19_pandemic, 2020. x[Online, accessed June 30, 2020].

